# Misfolded alpha synuclein co-occurrence with Alzheimer’s disease proteinopathy

**DOI:** 10.1101/2024.10.11.24315349

**Authors:** Erin M. Jonaitis, Karen MacLeod, Jennifer Lamoureux, Beckie Jeffers, Rachel L. Studer, John Middleton, Rachael E. Wilson, Nathaniel A. Chin, Ozioma C. Okonkwo, Barbara B. Bendlin, Sanjay Asthana, Cynthia M. Carlsson, Catherine L. Gallagher, Bruce Hermann, Sean McEvoy, Gwendlyn Kollmorgen, Henrik Zetterberg, Luis Concha-Marambio, Sterling C. Johnson, Russ M. Lebovitz, Rebecca E. Langhough

## Abstract

**INTRODUCTION:** Multi-etiology dementia necessitates in-vivo markers of copathologies including misfolded *α*-synuclein (syn). We measured misfolded syn aggregates (syn-seeds) via qualitative seed amplifcation assays (synSAA) and examined relationships with markers of Alzheimer’s disease (AD).

**METHODS:** Cerebrospinal fluid (CSF) was obtained from 420 participants in two Wisconsin AD risk cohorts (35% male; 91% cognitively unimpaired; mean (SD) age, 65.42 (7.78) years; education, 16.17 (2.23) years). synSAA results were compared to phosphorylated tau (T), beta amyloid (A), and clinical outcomes. Longitudinal cognition was modeled with mixed effects.

**RESULTS:** Syn positivity (synSAA+) co-occurred with T (in synSAA+ vs synSAA-, 36% vs 20% T+; p=0.011) and with cognitive impairment (10% vs 7% MCI; 10% vs 0% dementia; p=0.00050). synSAA+ participants’ cognitive performance declined ∼40% faster than synSAA-for Digit Symbol, but not other tests.

**DISCUSSION:** Findings support prevalent syn copathology in a mostly-unimpaired AD risk cohort. Future work will explore relationships with disease progression.

## 1 Introduction

Postmortem neuropathological investigations of patients with neurodegenerative disorders, including Alzheimer’s disease (AD), have revealed the coexistence of different proteinopathies. This profile, referred to as mixed dementia, may relate to patients’ presenting and evolving symptom complex, and may also have implications for treatment^1^. Identifying the underlying pathologies in persons with dementia or in those at-risk is of high priority, both to understand the biological disease processes and to select individuals for preventive and therapeutic clinical trials. Although much progress has been made in identifying biomarkers for beta-amyloid (A) and tau (T), tests for other proteinopathies have lagged.

Recently an *α*-synuclein (syn) seed amplification assay (synSAA) has been developed to identify individuals with underlying Lewy body disease (LBD) by detection of misfolded syn aggregates (syn-seeds) in cerebrospinal fluid (CSF)^2,3^. Reported concordance between synSAA results and neocortical neuropathology was high in two studies, with 97-100% of the cases with diffuse syn pathology detected by the assay (synSAA+), and 97-98.1% of the cases without syn pathology not detected (synSAA-)^4,5^. Other studies have shown that synSAA combined with other biomarker tests can be used to predict rate of decline in patients with and without AD co-pathology^6,7^. In fact, synSAA positivity has been shown to increase from MCI (A-T-) to AD dementia (A+T+), and to associate with the AD phenotypical presentation^5,8^.

In this study, we investigated the relationship between syn-seeds and concurrent AD biomarkers, clinical characteristics, and cognitive trajectories in predominantly unimpaired community-based cohorts enriched for AD risk. Specifically, we examined whether synSAA status was associated with elevated prevalence of concurrent CSF AD biomarker abnormalities or cognitive status, physical symptoms associated with Parkinson’s disease (PD) or LBD, or other concurrent characteristics. In addition, we examined whether syn-seeds, with or without comorbid AD biomarkers, were associated with faster cognitive decline.

## 2 Methods

### 2.1 Participants

Data were obtained and combined from participants in the Wisconsin Alzheimer’s Disease Research Center (WADRC), a community-based longitudinal cohort of individuals across the AD continuum^9^, and the Wisconsin Registry for Alzheimer’s Prevention (WRAP), a similar community cohort of individuals who were non-demented at baseline^10^. Both cohorts are enriched by design for a positive parental family history of AD. Participants were eligible for inclusion if they had had a lumbar puncture (LP) collected under a standard centerwide LP protocol, between December 5, 2018, and June 30, 2022, had consented to sample reuse, and had adequate sample available for the present analysis.

### 2.2 CSF collection

CSF collection is described in detail elsewhere^11^. Briefly, CSF was obtained via gentle extraction using a Sprotte 24- or 25-gauge atraumatic spinal needle. Samples were gently mixed and centrifuged at 2000 × g for 10 minutes at 4^∘^C within 30 minutes of extraction. After centrifugation, samples were aliquoted and stored in low-binding 0.5 mL tubes at -80^∘^C.

### 2.3 *α*-synuclein seed amplification assay

CSF samples were analyzed by the Amprion Clinical Laboratory using a rapid qualitative synSAA validated for clinical use under CLIA/CAP certifications. Each blinded sample was analyzed in triplicate (40*μ*L CSF per well) in a 96-well plate with a final reaction volume of 100*μ*L. The reaction mixture consisted of 0.3mg/mL rec-syn in 100mM PIPES pH 6.50, 0.44 M NaCl, 10*μ*M ThT, and 0.1% Sarkosyl, with two 1/8-inch silicon nitride beads per well. Plates were sealed using an Optical Adhesive Film, placed in a BMG LABTECH FLUOStar *Ω* Microplate Reader, and incubated at 42^∘^C with intermittent shaking (800 rpm orbital shaking for 1 min followed by 14 minutes of rest). Fluorescence readings (excitation wavelength, 440 nm; emission wavelength, 490 nm) were performed after every shaking cycle. After 20 hours of shaking/incubation, the maximum relative fluorescence (RFU; Fmax) of each well was determined, and CSF samples were classified based on a pre-established proprietary algorithm^8^. Based on this algorithm, if all three replicates return a positive result, the sample is classified as Detected; if 0 or 1 replicates return a positive result, the sample is classified as Not Detected; and if 2 replicates return a positive result the sample is classified as Indeterminate. The Detected category is further subdivided on the basis of Fmax into Detected-1 (syn seeding aggregates detected; amplification profile consistent with that found predominantly in subjects with neuronal synuclein disease) and Detected-2 (syn seeding aggregates detected; amplification profile consistent with that found predominantly in patients with multiple system atrophy). This clinical version of the assay was performed according to standard operating procedures in accordance with CLIA regulations.

### 2.4 Alzheimer’s disease and NeuroToolKit biomarker assays

Alzheimer’s disease biomarkers were assayed at the Clinical Neurochemistry Laboratory in Gothenburg, Sweden, using fully automated electrochemiluminescent immunoassays. CSF levels of amyloid beta-42 (A*β*_42_) and phosphorylated tau-181 (pTau_181_) were measured using the in vitro diagnostic (IVD) Elecsys® β-amyloid(1-42) CSF (second-generation) and Phospho-Tau(181P) CSF assays, and amyloid beta-40 (A*β*_40_) levels were measured using the robust prototype assay Elecsys® β-amyloid(1-40). CSF levels of neurofilament light-chain (NfL), glial fibrillary acidic protein (GFAP), neurogranin (Ng), soluble triggering receptor expressed on myeloid cells (sTREM2), and chitinase-3-like protein 1 (YKL40) were measured using the NeuroToolKit (NTK), a panel of exploratory robust prototype assays, on a Cobas^®^ e 411 analyzer (Roche Diagnostics International Ltd). Derivation of cutpoints for Alzheimer’s disease biomarker positivity has been described elsewhere^12^. Briefly, thresholds for two ratios, A*β*_42/40_ and pTau_181_/A*β*_42_, were determined by fitting ROC curves against an amyloid PET criterion. The first of these ratios is often used as indicator of amyloid positivity; the second has been variously construed as an amyloid positivity marker^12^ or as a single-dimensional marker of overall AD pathology^13^. The threshold for pTau was defined at the upper bound of a 95% confidence interval on the distribution of pTau_181_ levels in a subset of young, cognitively unimpaired, CSF amyloid-negative participants.

### 2.5 Cognitive assessment

Participants in each cohort complete neuropsychological tests every 1-2 years. The WRAP battery has been described in detail elsewhere^10^. The WADRC battery includes several tests that are core to the WRAP battery, as well as those specified in the National Alzheimer’s Coordinating Center Uniform Data Sets^14^. For the present study, we used three cognitive tests that measure aspects of executive function, including visuomotor speed: the Trail-Making Test, parts A and B, and their difference^15^, Wechsler Adult Intelligence Scale-Digit Span Backward^16,17^ and Digit Symbol Substitution^16^. We also included a cognitive composite that is sensitive to cognitive decline associated with Alzheimer’s disease, a three-test Preclinical Alzheimer’s Cognitive Composite^18,19^ comprising the sum of learning trials on the Rey Auditory Verbal Learning test^20^, delayed performance on a story recall task^14,21^, and Letter Fluency from the COWAT^22^. Scores were equated using published crosswalks where available, and internal equipercentile mappings otherwise^23,24^.

Cognitive status was assigned at each neuropsychological assessment via a multidisciplinary clinical consensus conference. Consensus review procedures are similar across cohorts; however, due to the high prevalence of cognitively unimpaired individuals in the WRAP cohort, the WRAP consensus process begins with an algorithmic screening in which only those who fall at least 1.5 standard deviations below robust internal norms and those whose raw test scores or study partner questionnaire responses exceed thresholds for concern are reviewed in detail. The remainder are assigned a status indicating no clinically significant impairment^25^.

### 2.6 Self-reported health variables

Along with cognitive assessment, participants complete detailed health questionnaires every 1-2 years. Participant physical activity was estimated in MET-hours per week based on responses to questions about mild, moderate, and vigorous activity and walking outside the home. Recent-onset depression was defined as any first report of depression at a visit after baseline and within five years of lumbar puncture.

### 2.7 Clinical evaluation

Clinical symptoms were assessed by a nurse practitioner following the standard procedures associated with the National Alzheimer’s Coordinating Center Uniform Data Set version 3, forms B8 and B9. Presence of a symptom at any visit was counted as a positive sign.

### 2.8 Statistical methods

Primary analyses examined relationships between binary synSAA status and other variables. synSAA results of Detected-1 were coded as synSAA+, Not Detected as synSAA-, and both Detected-2 and Indeterminate as missing values. The co-occurrence in CSF of syn-seeds (based on positive/negative status as detected via synSAA) with, variously, AD biomarkers (based on positive/negative status for amyloid beta, A*β*_42/40_ and pTau_181_/A*β*_42_, and phosphorylated tau, pTau_181_) and clinical symptoms (any neurological finding; Parkinsonian signs; REM behavior disorder; recent-onset depression; clinically-significant cognitive impairment as adjudicated by consensus conference) was examined with chi-square tests. Confidence intervals for proportions were estimated using Agresti-Coull intervals^26^. Group differences in cognition and exercise were examined with t-tests and Mann-Whitney U tests, respectively. For these analyses, missing data were excluded on a pairwise basis. Sensitivity analyses assessed the dependence of our findings on age. Chi-square tests were repeated in subsets stratified by age into younger (<65) and older (>=65). In addition, we used logistic regression to model binary synSAA (synSAA+=1) as a function of these biomarker and clinical predictors after controlling for age.

Exploratory analyses compared levels of NTK biomarkers between groups defined on the basis of joint synSAA status and T status. Continuous biomarker outcomes were modeled using separate one-way ANOVAs (synSAA-/T-; synSAA-/T+; synSAA+/T-; synSAA+/T+). Initial examination revealed an extreme outlier on NfL (result > 35 times the median result); this datapoint was removed for a post-hoc analysis.

Cognitive trajectories were estimated using linear mixed effects models of Trail-Making Test B, the Trail-Making Test difference score (B - A), Backward Digit Span, Digit Symbol Substitution, and a three-test Preclinical Alzheimer’s Cognitive Composite (PACC-3). All models included subject-level random intercepts and age slopes with unstructured covariance. The fixed effects of age were modeled by centering age and including up to a quadratic term. Covariates included sex, education, and number of prior cognitive assessments per protocol (to adjust for practice effects). For each outcome, we examined whether last known synSAA status modified age trajectories (models 1A-4A). In a second set of models (1B-4B), we further examined syn pathology effects after adjusting for separate age interactions with binarized A*β*_42/40_ and pTau_181_ statuses. Finally, an exploratory set of models (1C-4C) used pTau_181_/A*β*_42_ as a single-dimensional indicator of AD pathology to ask whether AD biomarker status and synSAA status interact with age or each other to predict cognition. For this analysis, pairwise contrasts of simple slopes in all four biomarker groups were estimated using Tukey’s adjustment for multiple comparisons. In all three sets of models, nonsignificant interaction terms were removed (p>.1). Biomarker negative status was the reference category for each marker. For these analyses, participants missing data on any predictor were excluded on a listwise basis to facilitate nested model comparisons within outcomes (i.e., comparisons of models A and B for a given outcome). However, all participants with at least one observation on any outcome were included in analyses of that outcome.

Statistical analyses were conducted in R^27^. Mixed effects models were fit with the lmerTest package^28^ and marginal effects were estimated using the packages ggeffects^29^ and emmeans^30^.

## 3 Results

### 3.1 Participants

543 LPs were performed during the relevant period, of which 515 samples on 420 participants had sufficient volume remaining for synSAA analysis. This included 214 participants from WADRC (at visit closest to LP, 183 CU; 19 MCI; 6 Dementia) and 206 participants from WRAP (195 CU; 11 MCI; 0 Dementia). Two participants had a synSAA result of Indeterminate, and one had a result of Detected-2, and were excluded from further analyses. Among those with synSAA results meeting criteria for inclusion, 12% were identified as synSAA+. Participant characteristics for those with includable synSAA results are shown in Table 1, overall and by synSAA status. The overall sample had more women than men (35% male), but the gender balance differed by synSAA status (synSAA+, 49% male; synSAA-, 33% male). Further, synSAA+ participants tended to be older at LP (synSAA+, 69.04 (7.47); synSAA-, 64.93 (7.70)). No differences in education or APOE genotype were seen (p>.05).

**Table 1.** Sample characteristics of participants, overall and by synSAA status. Three participants were excluded (N=1 with results suggesting multiple system atrophy; N=2 with indeterminate results). A*β*_42/40_, positive amyloid beta 42/40 status; APOE4 = carriage of at least one apolipoprotein $$4 allele; CU, cognitively unimpaired; LP, lumbar puncture; MCI, mild cognitive impairment; MET-hours, metabolic equivalent hours per week; Neurological findings, presence of any sign or symptom suggestive of (a) age-related change or (b) a dementing disorder; pTau_181_, positive phosphorylated tau-181 status; pTau_181_/A*β*_42_, positive pTau to amyloid ratio; REM, rapid eye movement; synSAA, *α*-synuclein seed amplification assay status; URG, underrepresented racial group membership.

| Variable | Overall | synSAA- | synSAA+ | p |
| --- | --- | --- | --- | --- |
| N | 417 | 368/417 (88%; 85%-91%) | 49/417 (12%; 9%-15%) | – |
| <u>Demographics</u> |  |  |  |  |
| Gender, male, N (%) | 146/417 (35%; 31%-40%) | 122/368 (33%; 29%-38%) | 24/49 (49%; 36%-63%) | 0.034 |
| URG member, Yes, N (%) | 48/417 (12%; 9%-15%) | 46/368 (12%; 9%-16%) | 2/49 (4%; 0%-14%) | 0.099 |
| Education, years, mean (SD) | 16.17 (2.23) | 16.14 (2.21) | 16.39 (2.39) | 0.50 |
| APOE $\epsilon 4$+, N (%) | 164/360 (46%; 40%-51%) | 140/315 (44%; 39%-50%) | 24/45 (53%; 39%-67%) | 0.34 |
| Age at LP, years, mean (SD) | 65.42 (7.78) | 64.93 (7.70) | 69.04 (7.47) | 0.00064 |
| Time between LP and cognition, years, mean (SD) | 0.026 (0.49) | 0.030 (0.52) | 0.0012 (0.0033) | 0.30 |
| <u>AD Biomarkers</u> |  |  |  |  |
| $A\beta_{42/40}$ positive, N (%) | 127/412 (31%; 27%-35%) | 108/364 (30%; 25%-35%) | 19/48 (40%; 27%-54%) | 0.17 |
| pTau<sub>181</sub>/$A\beta_{42}$ positive, N (%) | 118/405 (29%; 25%-34%) | 99/359 (28%; 23%-32%) | 19/46 (41%; 28%-56%) | 0.067 |
| pTau<sub>181</sub> positive, N (%) | 88/410 (21%; 18%-26%) | 71/363 (20%; 16%-24%) | 17/47 (36%; 24%-51%) | 0.011 |
| <u>Cognitive status at nearest assessment</u> |  |  |  | 0.00050 |
| Cognitive status, CU, N (%) | 375/411 (91%; 88%-94%) | 336/362 (93%; 90%-95%) | 39/49 (80%; 66%-89%) |  |
| Cognitive status, MCI, N (%) | 30/411 (7%; 5%-10%) | 25/362 (7%; 5%-10%) | 5/49 (10%; 4%-22%) |  |

| **Variable** | **Overall** | **synSAA-** | **synSAA+** | **p** |
| --- | --- | --- | --- | --- |
| Cognitive status, Dementia, N (%) | 6/411 (1%; 1%-3%) | 1/362 (0%; 0%-2%) | 5/49 (10%; 4%-22%) |  |
| <u>Neurological findings</u> |  |  |  | 0.018 |
| Neurological findings (age-related), N (%) | 72/412 (17%; 14%-21%) | 64/365 (18%; 14%-22%) | 8/47 (17%; 9%-30%) |  |
| Neurological findings (dementia-related), N (%) | 29/412 (7%; 5%-10%) | 21/365 (6%; 4%-9%) | 8/47 (17%; 9%-30%) |  |
| Parkinsonian signs, N (%) | 22/417 (5%; 3%-8%) | 17/368 (5%; 3%-7%) | 5/49 (10%; 4%-22%) | 0.16 |
| REM behavior disorder, N (%) | 6/417 (1%; 1%-3%) | 4/368 (1%; 0%-3%) | 2/49 (4%; 0%-14%) | 0.15 |
| <u>Other health variables</u> |  |  |  |  |
| New-onset depression, N (%) | 13/417 (3%; 2%-5%) | 10/368 (3%; 1%-5%) | 3/49 (6%; 1%-17%) | 0.40 |
| Exercise, MET-hours per week, mean (SD) | 15.75 (0-83.00) | 16.00 (0-83.00) | 12.00 (0-39.75) | 0.099 |

### 3.2 synSAA and AD biomarkers

Among the 417 participants observed with includable results for synSAA, 23 (6%; 95% CI, 4%-8%) were positive for syn-seeds and any of the three AD CSF markers; 26 (6%; 4%-9%) were positive for syn-seeds only, and not any of the AD markers; 132 (32%; 27%-36%) were positive for any of the AD biomarkers, but negative for syn-seeds; and 236 (57%; 52%-61%) were negative for all four biomarkers. Among 92 participants with more than one synSAA analysis, none were discordant, suggesting medium-term stability of this measure (range of inter-LP intervals observed: 0-3.1 years).

synSAA status was associated with pTau_181_ status (p=0.011), and weakly with pTau_181_/A*β*_42_ status (p=0.067), but not with A*β*_42/40_ status (p=0.17). Among pTau_181_+ participants, 17/88 (19%; 12%-29%) were also synSAA+, compared to 30/322 (9%; 7%-13%) pTau_181_- participants. For pTau_181_/A*β*_42_+ and pTau_181_/A*β*_42_-, the figures were 19/118 (16%; 10%-24%) and 27/287 (9%; 7%-13%), respectively. However, sensitivity analyses suggest that these relationships between synSAA and other biomarkers may be affected by age: after stratifying into older and younger subgroups, chi square tests are no longer significant in either group (all p>0.1). The reduction of N and resulting loss of power may have influenced this shift; nevertheless, logistic regression models controlling for age produce similar findings, with no p-values reaching statistical significance. P-values for primary and sensitivity analyses are in Supplementary Table 1; a forest plot of overall and age-stratified confidence intervals for synSAA status by AD biomarker group is shown in Supplementary Figure 1.

### 3.3 synSAA and exploratory biomarkers

All markers except for NfL exhibited differences between some pair of groups (p<.0001). GFAP levels were lower in synSAA-/T-participants than in either of the T+ groups. Ng and YKL40 levels were lower in all T-groups than in all T+ groups, irrespective of synSAA status. sTREM2 levels were lower in synSAA-/T-participants than in either of the T+ groups, and were lower in synSAA+/T-participants than in synSAA+/T+ participants. Examination of the NfL data revealed that one NfL observation had a value more than 35 times as high as the median; a post-hoc comparison removing this outlier revealed a pattern of group means for NfL similar to Ng and YKL40. Group comparisons excluding the outlier are shown in Table 2 and Figure 1.

**Figure 1:**
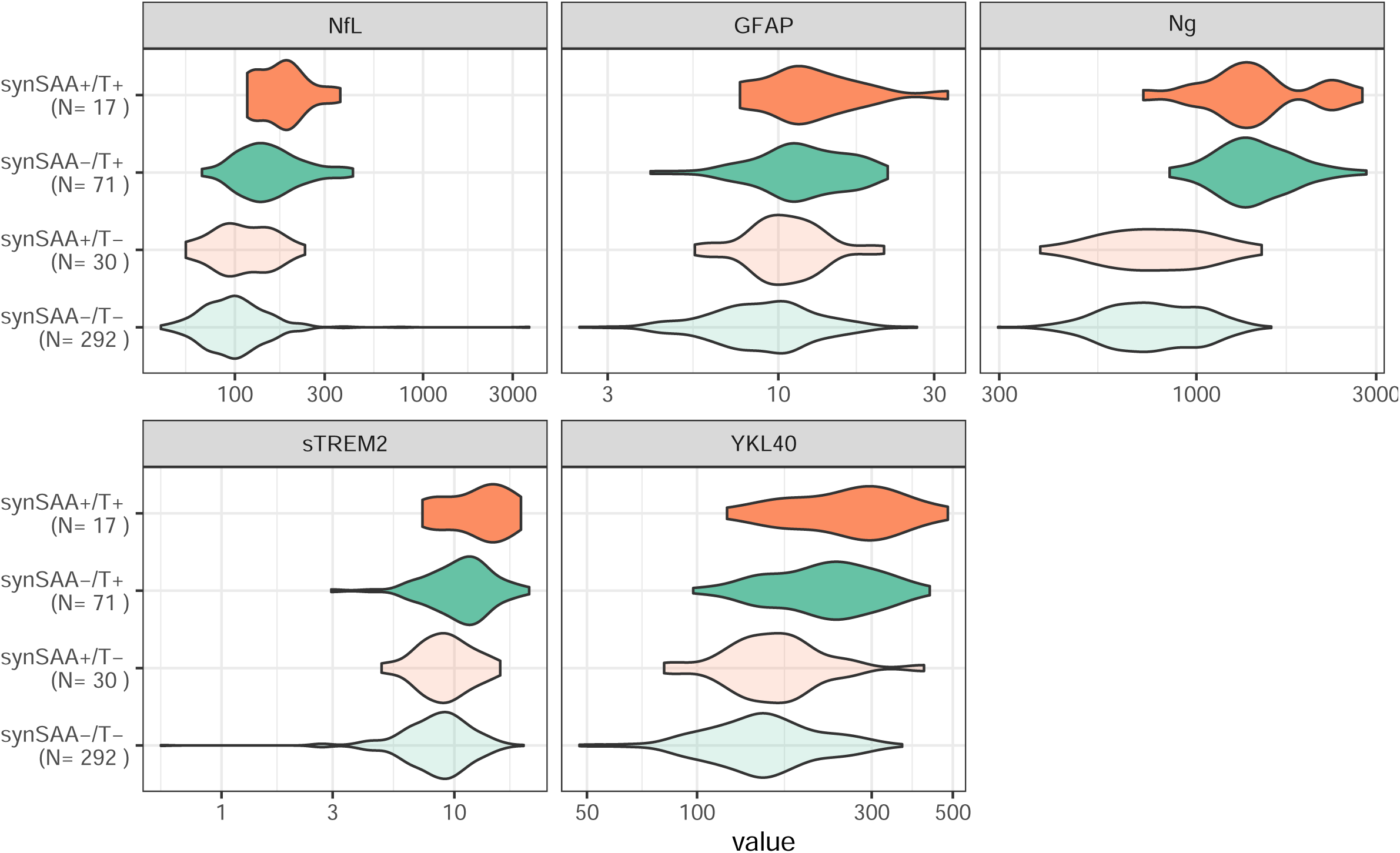
Violin plots of robust prototype biomarkers from the NeuroToolKit in groups defined by synSAA status and T status. One NfL observation whose value was more than 35 times the median value was removed to aid in visualization. GFAP, glial fibrillary acidic protein; NfL, neurofilament light-chain; Ng, neurogranin; sTREM2, soluble triggering receptor expressed on myeloid cells 2; synSAA, α-synuclein seed amplification assay status; T, phosphorylated tau 181 status; YKL40, chitinase-3-like protein 1.

**Table 2.** Comparison of robust prototype biomarkers from the NeuroToolKit in groups defined by synSAA status and T status. One NfL observation whose value was more than 35 times the median value was removed for comparisons; if included, NfL comparisons are non-significant, p>.10. GFAP, glial fibrillary acidic protein; NfL, neurofilament light-chain; Ng, neurogranin; sTREM2, soluble triggering receptor expressed on myeloid cells 2; synSAA, *α*-synuclein seed amplification assay status; T, phosphorylated tau 181 status; YKL40, chitinase-3-like protein 1.

| Variable | synSAA-/T- | synSAA+/T- | synSAA-/T+ | synSAA+/T+ | p | Contrasts |
| --- | --- | --- | --- | --- | --- | --- |
| NfL | 120.45 (216.74) | 117.30 (44.02) | 166.42 (77.44) | 187.49 (66.97) | <.0001 | synSAA-/T- < synSAA-/T+;<br>synSAA-/T- < synSAA+/T+;<br>synSAA+/T- < synSAA-/T+;<br>synSAA+/T- < synSAA+/T+ |
| GFAP | 9.50 (3.55) | 10.86 (3.21) | 12.38 (3.89) | 13.60 (6.12) | <.0001 | synSAA-/T- < synSAA-/T+;<br>synSAA-/T- < synSAA+/T+; |
| Ng | 798.81 (229.94) | 822.77 (268.41) | 1466.87 (375.48) | 1522.33 (549.18) | <.0001 | synSAA-/T- < synSAA-/T+;<br>synSAA-/T- < synSAA+/T+;<br>synSAA+/T- < synSAA-/T+;<br>synSAA+/T- < synSAA+/T+ |
| sTREM2 | 8.79 (2.78) | 9.42 (2.56) | 11.06 (3.41) | 12.96 (3.88) | <.0001 | synSAA-/T- < synSAA-/T+;<br>synSAA-/T- < synSAA+/T+;<br>synSAA+/T- < synSAA+/T+ |
| YKL40 | 159.85 (56.43) | 171.38 (63.45) | 233.55 (75.09) | 269.47 (93.81) | <.0001 | synSAA-/T- < synSAA-/T+;<br>synSAA-/T- < synSAA+/T+;<br>synSAA+/T- < synSAA-/T+;<br>synSAA+/T- < synSAA+/T+ |

### 3.4 synSAA, clinical impairment, and health

Compared to synSAA-participants, synSAA+ had a higher incidence of cognitive impairment as judged by consensus conference (p=0.00050; synSAA+, MCI=5/49 (10%; 4%-22%), Dementia=5/49 (10%; 4%-22%); synSAA-, MCI=25/362 (7%; 5%-10%), Dementia=1/362 (0%; 0%-2%)). In the age-stratified sensitivity analysis, however, this relationship was not seen in the younger group (Supp. Table 1).

synSAA+ participants also exhibited more neurological findings (p=0.018). However, drilling down, we did not see similar evidence linking syn-seeds to excess Parkinsonian signs (p=0.16), REM behavior disorder (p=0.15), or depression (p=0.40). Age-stratified sensitivity analyses indicated that within the younger group, synSAA results were weakly associated with both neurological findings and depression, but these relationships were not seen in the older group. When analyzed instead with logistic regression models controlling for age, no relationships were seen between syn-seeds and any neurological predictor. P-values for primary and sensitivity analyses are in Supplementary Table 1; a forest plot of overall and age-stratified confidence intervals for synSAA status by AD biomarker group is shown in Supplementary Figure 1.

Self-reported physical activity did not differ significantly between synSAA+ and synSAA-participants (synSAA+, median (range)=12.00 (0-39.75); synSAA-, median (range)=16.00 (0-83.00); p=0.099).

### 3.5 Syn-seeds and cognitive trajectories

Availability of cognitive follow-up depended on the outcome, with the least for Trails Difference (N_*sub*_=387; N_*obs*_=2675) and Digit Symbol (N_*sub*_=388; N_*obs*_=2079), and the most for Digit Span Backward (N_*sub*_=403; N_*obs*_=2972). Cognitive statuses at baseline were as follows: CU, 375; MCI, 22; Dementia, 4).

Full results of cognitive trajectory analyses are shown in Tables 3 and 4 by each of the five cognitive outcomes and two model sets (Table 3, model A set: covariates + age + age^2^ + synSAA + (synSAA × age) + (synSAA × age^2^); Table 4, model B set: covariates + age + age^2^ + synSAA + (synSAA × age) + (synSAA × age^2^) + A*β*_42/40_ + (A*β*_42/40_ × age) + (A*β*_42/40_ × age^2^) + pTau_181_ + (pTau_181_ × age) + (pTau_181_ × age^2^). Simple age slopes for synSAA- and synSAA+ estimated from the model B sets are depicted for each outcome in Figure 2, paneling each figure by A/T statuses.

**Figure 2:**
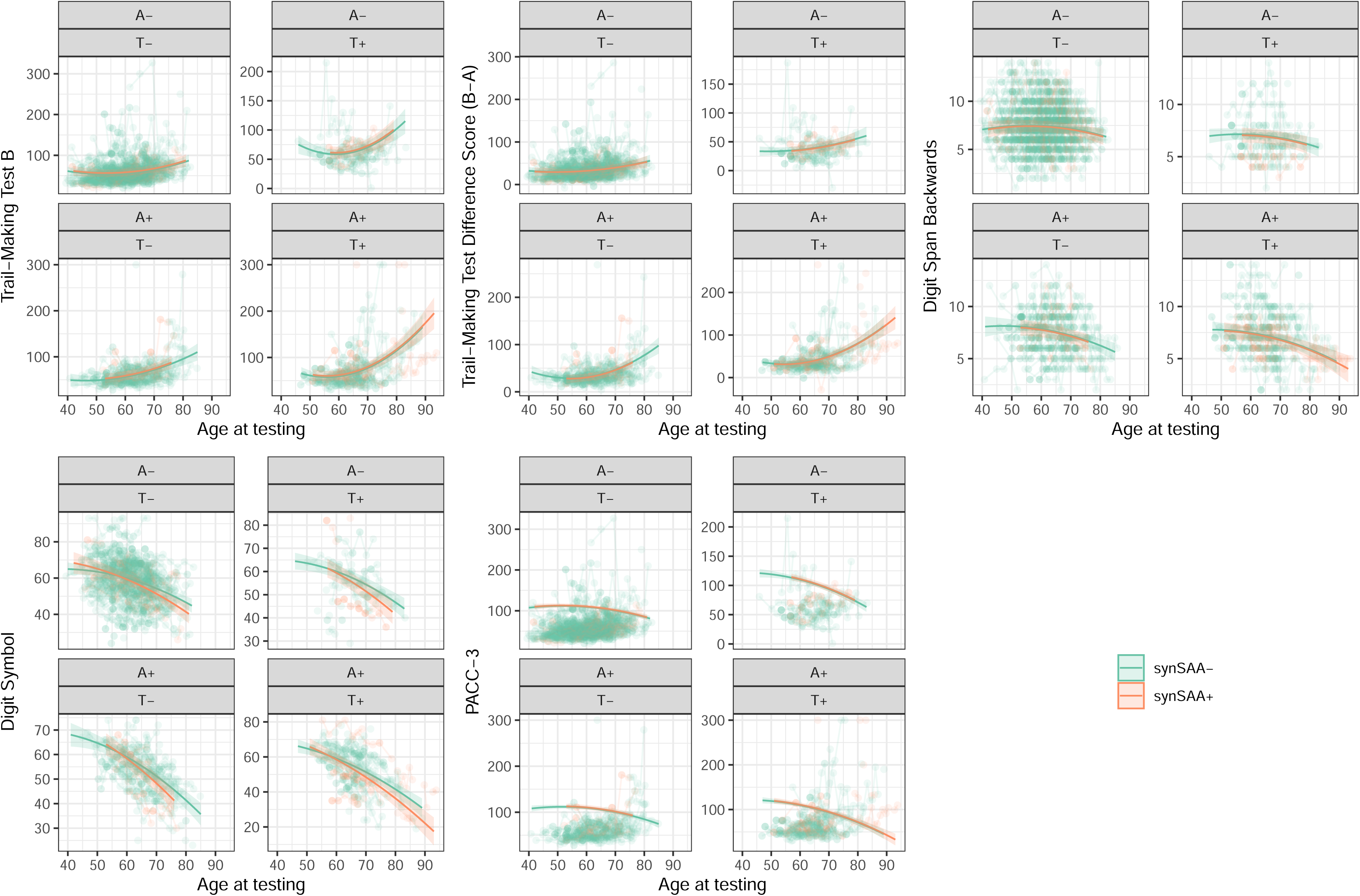
Results of nested linear mixed effects models of cognitive tests associated with executive function (Trail-Making Test B; Trail-Making Test B-A Difference Score; Digit Span Backward; Digit Symbol Substitution Test) and a global preclinical Alzheimer’s cognitive composite (PACC-3). Model-predicted values and confidence bands derived from final models represented in Table 2. Predictors not shown directly in the graph have been set to their average value. The largest model examined the effect of binary synSAA, synSAA × age (centered at 60), and synSAA × age^2^, controlling for sex, education, and prior exposure to the battery, alongside binary A*β*_42/40_ and pTau_181_ and their interactions with age and age^2^. From this largest model, nonsignificant interaction terms (p>.1) were removed. The spaghetti plot layer beneath represents individual participants’ measurements over time.

**Table 3.**
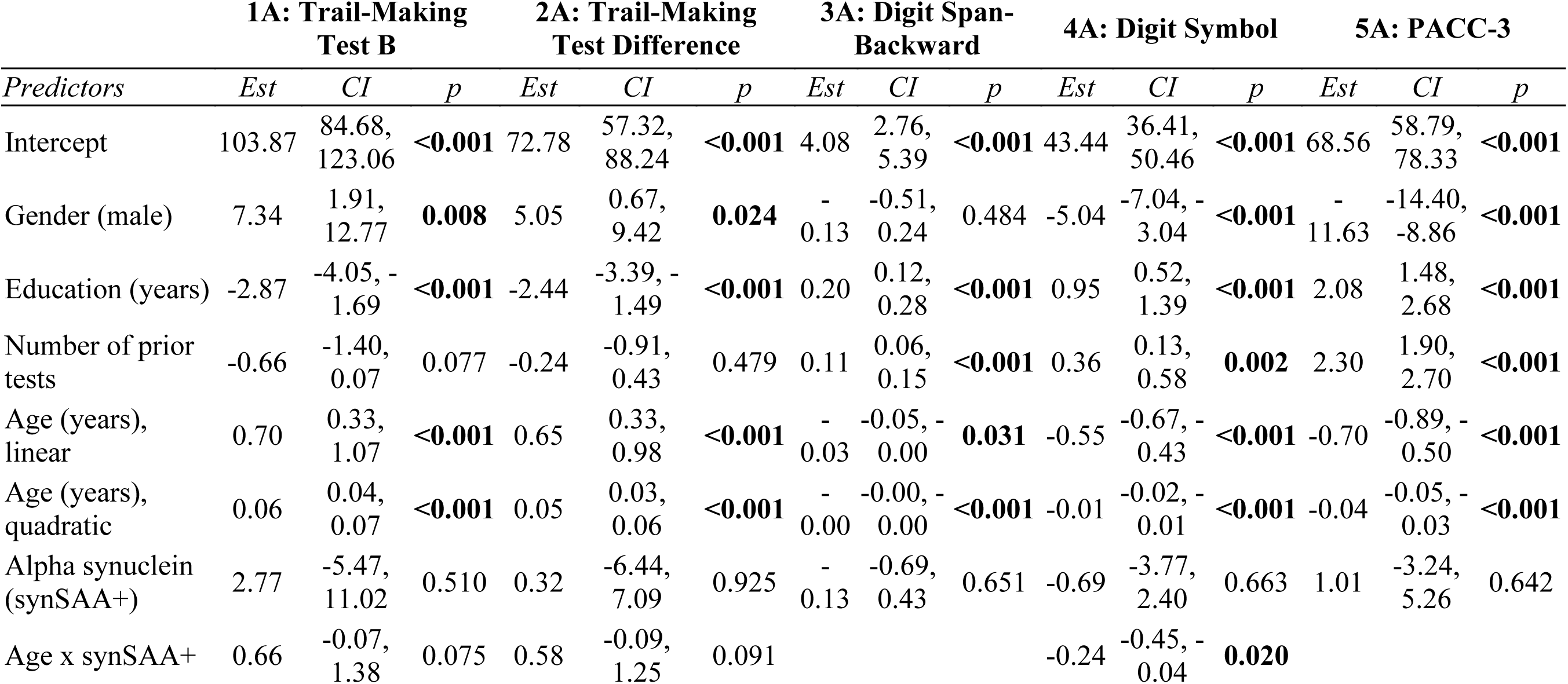
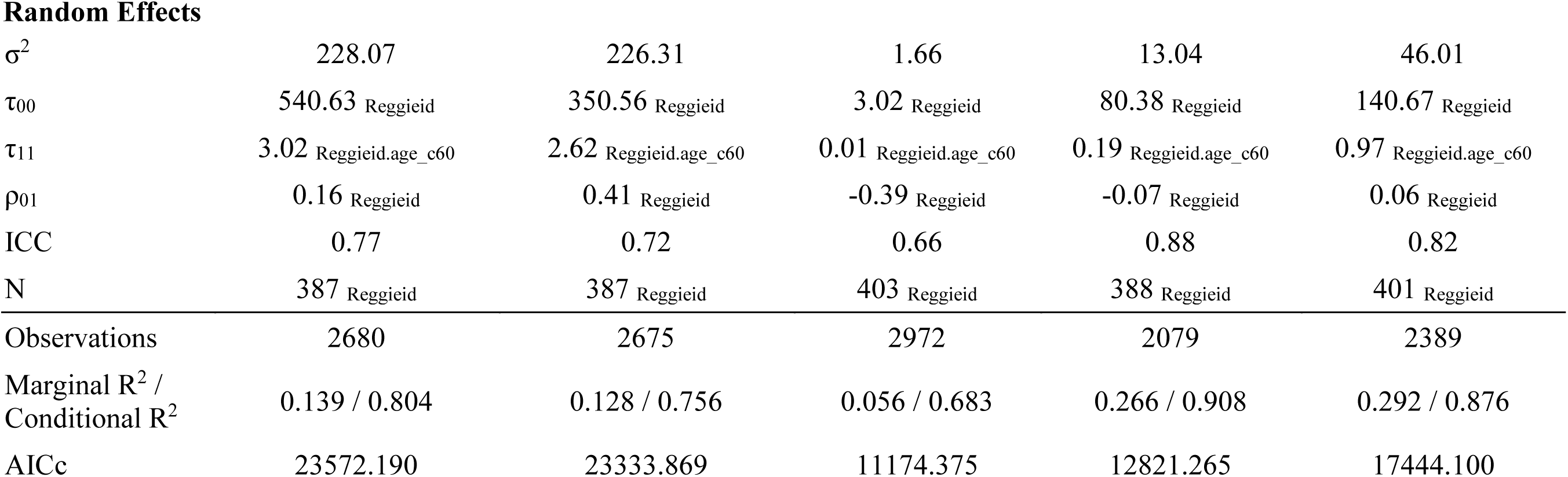
Results of nested linear mixed effects models of cognitive tests associated with executive function (Trail-Making Test B; Trail-Making Test B-A Difference Score; Digit Span Backward; Digit Symbol Substitution Test) and a global preclinical Alzheimer’s cognitive composite (PACC-3). Model set A examined the effect of binary synSAA, synSAA × age (centered at 60), and synSAA × age^2^, controlling for sex, education, and prior exposure to the battery. Nonsignificant interaction terms (p>.1) were removed.

|  | **1A: Trail-Making**<br>**Test B** |  |  | **2A: Trail-Making**<br>**Test Difference** |  |  | **3A: Digit Span-**<br>**Backward** |  |  | **4A: Digit Symbol** |  |  | **5A: PACC-3** |  |  |
| --- | --- | --- | --- | --- | --- | --- | --- | --- | --- | --- | --- | --- | --- | --- | --- |
| *Predictors* | *Est* | *CI* | *p* | *Est* | *CI* | *p* | *Est* | *CI* | *p* | *Est* | *CI* | *p* | *Est* | *CI* | *p* |
| Intercept | 103.87 | 84.68,<br>123.06 | **<0.001** | 72.78 | 57.32,<br>88.24 | **<0.001** | 4.08 | 2.76,<br>5.39 | **<0.001** | 43.44 | 36.41,<br>50.46 | **<0.001** | 68.56 | 58.79,<br>78.33 | **<0.001** |
| Gender (male) | 7.34 | 1.91,<br>12.77 | **0.008** | 5.05 | 0.67,<br>9.42 | **0.024** | -<br>0.13 | -0.51,<br>0.24 | 0.484 | -5.04 | -7.04, -<br>3.04 | **<0.001** | -<br>11.63 | -14.40,<br>-8.86 | **<0.001** |
| Education (years) | -2.87 | -4.05, -<br>1.69 | **<0.001** | -2.44 | -3.39, -<br>1.49 | **<0.001** | 0.20 | 0.12,<br>0.28 | **<0.001** | 0.95 | 0.52,<br>1.39 | **<0.001** | 2.08 | 1.48,<br>2.68 | **<0.001** |
| Number of prior<br>tests | -0.66 | -1.40,<br>0.07 | 0.077 | -0.24 | -0.91,<br>0.43 | 0.479 | 0.11 | 0.06,<br>0.15 | **<0.001** | 0.36 | 0.13,<br>0.58 | **0.002** | 2.30 | 1.90,<br>2.70 | **<0.001** |
| Age (years),<br>linear | 0.70 | 0.33,<br>1.07 | **<0.001** | 0.65 | 0.33,<br>0.98 | **<0.001** | -<br>0.03 | -0.05, -<br>0.00 | **0.031** | -0.55 | -0.67, -<br>0.43 | **<0.001** | -0.70 | -0.89, -<br>0.50 | **<0.001** |
| Age (years),<br>quadratic | 0.06 | 0.04,<br>0.07 | **<0.001** | 0.05 | 0.03,<br>0.06 | **<0.001** | -<br>0.00 | -0.00, -<br>0.00 | **<0.001** | -0.01 | -0.02, -<br>0.01 | **<0.001** | -0.04 | -0.05, -<br>0.03 | **<0.001** |
| Alpha synuclein<br>(synSAA+) | 2.77 | -5.47,<br>11.02 | 0.510 | 0.32 | -6.44,<br>7.09 | 0.925 | -<br>0.13 | -0.69,<br>0.43 | 0.651 | -0.69 | -3.77,<br>2.40 | 0.663 | 1.01 | -3.24,<br>5.26 | 0.642 |
| Age x synSAA+ | 0.66 | -0.07,<br>1.38 | 0.075 | 0.58 | -0.09,<br>1.25 | 0.091 |  |  |  | -0.24 | -0.45, -<br>0.04 | **0.020** |  |  |  |

**Random Effects**
|  |  |  |  |  |  |
| --- | --- | --- | --- | --- | --- |
| $\sigma^2$ | 228.07 | 226.31 | 1.66 | 13.04 | 46.01 |
| $\tau_{00}$ | 540.63 Reggieid | 350.56 Reggieid | 3.02 Reggieid | 80.38 Reggieid | 140.67 Reggieid |
| $\tau_{11}$ | 3.02 Reggieid.age c60 | 2.62 Reggieid.age c60 | 0.01 Reggieid.age c60 | 0.19 Reggieid.age c60 | 0.97 Reggieid.age c60 |
| $\rho_{01}$ | 0.16 Reggieid | 0.41 Reggieid | -0.39 Reggieid | -0.07 Reggieid | 0.06 Reggieid |
| ICC | 0.77 | 0.72 | 0.66 | 0.88 | 0.82 |
| N | 387 Reggieid | 387 Reggieid | 403 Reggieid | 388 Reggieid | 401 Reggieid |
| Observations | 2680 | 2675 | 2972 | 2079 | 2389 |
| Marginal $R^2$ /<br>Conditional $R^2$ | 0.139 / 0.804 | 0.128 / 0.756 | 0.056 / 0.683 | 0.266 / 0.908 | 0.292 / 0.876 |
| AICc | 23572.190 | 23333.869 | 11174.375 | 12821.265 | 17444.100 |

**Table 4.**
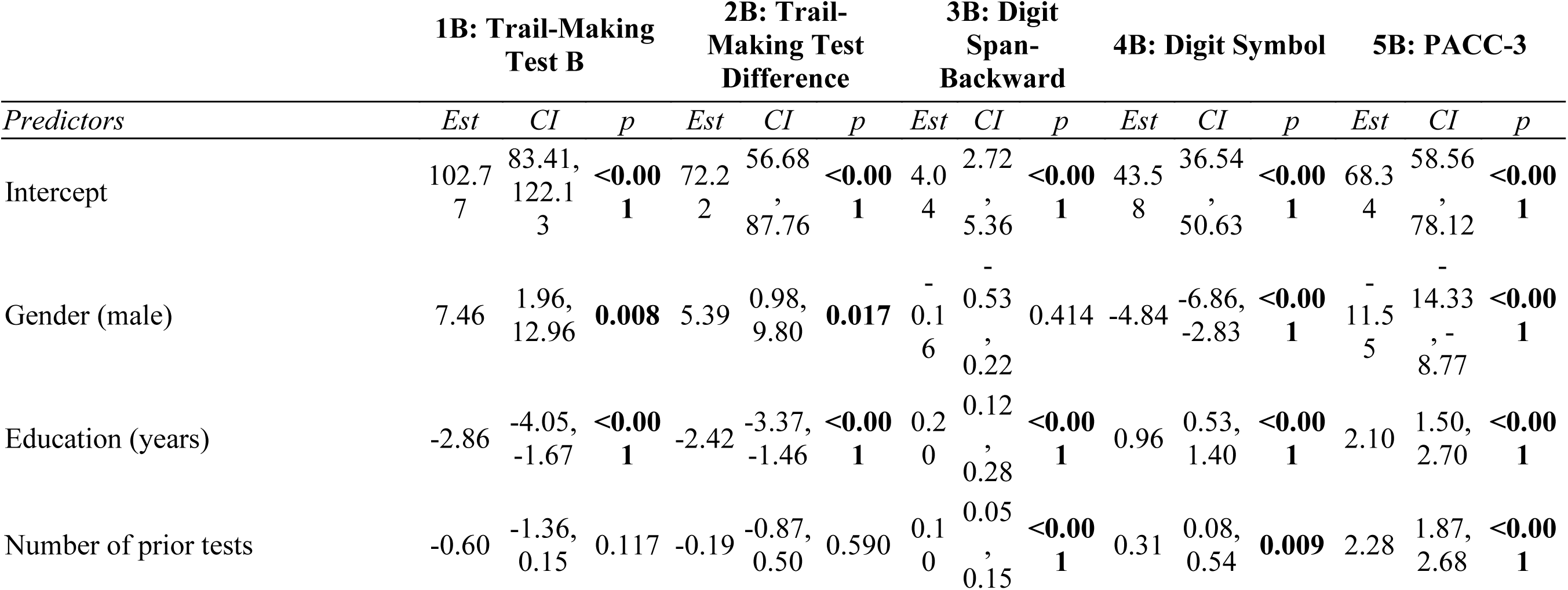
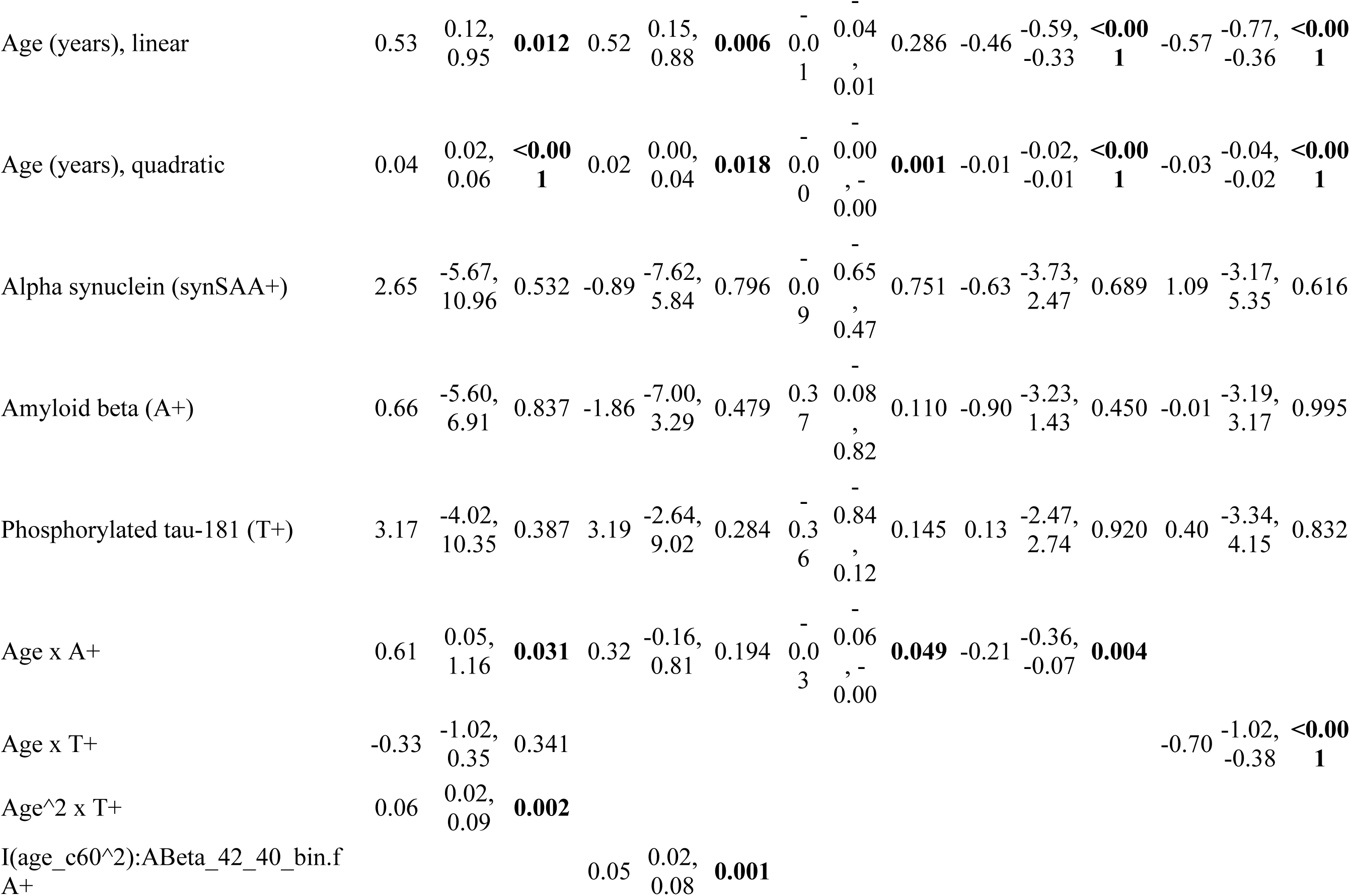
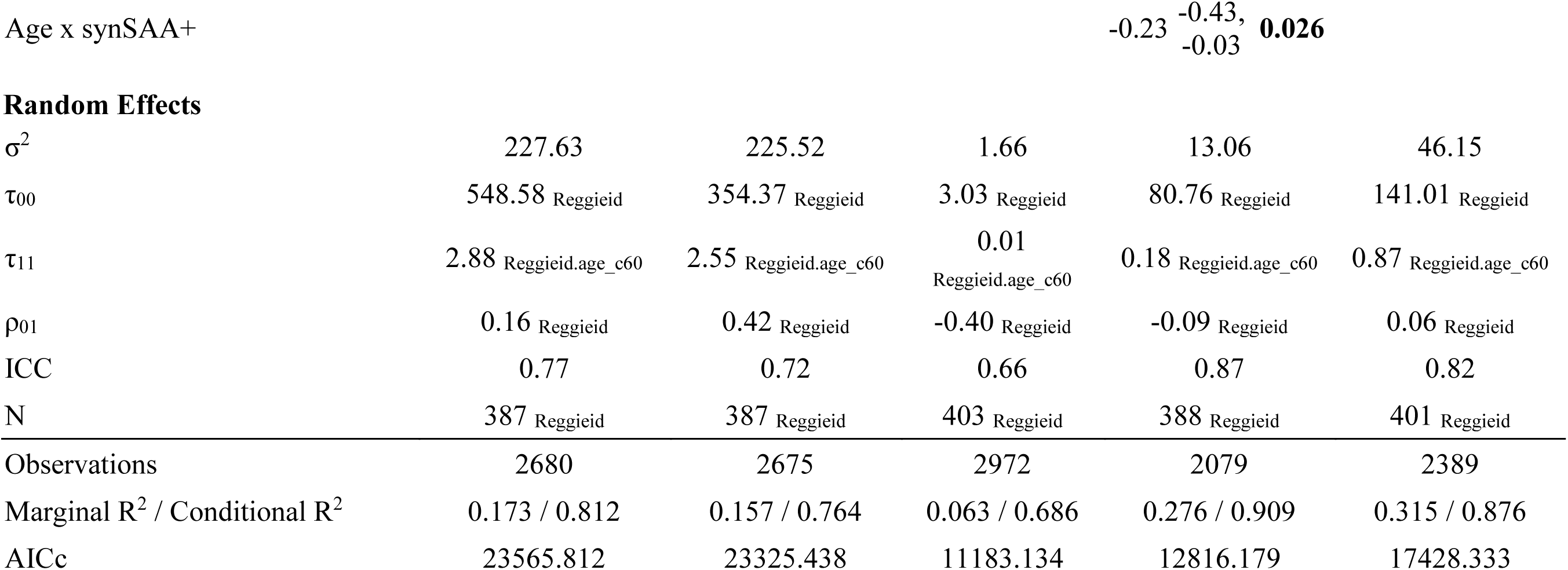
Results of nested linear mixed effects models of cognitive tests associated with executive function (Trail-Making Test B; Trail-Making Test B-A Difference Score; Digit Span Backward; Digit Symbol Substitution Test) and a global preclinical Alzheimer’s cognitive composite (PACC-3). Model set B examined the effect of binary synSAA, synSAA × age (centered at 60), and synSAA × age^2^, along with binary A*β*_42/40_ and pTau_181_ and their interactions with age and age^2^. As in Model set A, all models controlled for sex, education, and prior exposure to the battery. Nonsignificant interaction terms (p>.1) were removed.

#### Trails B

In model 1A, the interaction estimates marginally more age-related worsening in synSAA+ than synSAA- (*β*_synSAA×age_ = 0.66 seconds per year, p=0.076). However, once AD biomarkers and their interactions with age terms are added in model 1B, *β*_synSAA×age_ is attenuated (p>.1), while both A+ and T+ statuses are associated with significantly worse age-related change (*β*_A*β*42/40×age_ = 0.61, p=0.032; 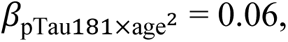 p=0.0021).

#### Trails Difference

In model 2A, the interaction estimates marginally more age-related worsening in synSAA+ than synSAA- (*β*_synSAA×age_ = 0.58 seconds per year, p=0.092). However, once AD biomarkers and their interactions with age terms are added in model 2B, *β*_synSAA×age_ is attenuated (p>.1), while both A+ status (but not T+ status) is associated with significantly greater acceleration in age-related change (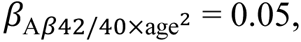 p=0.0010).

#### Digit Span-Backward

No significant differences in age-related change are seen by synSAA status in either model 3A or 3B. In model 3B, faster declines are seen for A+ vs A- (*β*_A*β*42/40×age_ = -0.03, p=0.050), but not for T+ vs T-.

#### Digit Symbol

In model 4A, faster age-related declines were observed in synSAA+ compared to synSAA- (*β*_synSAA×age_ = -0.24 points per year, p=0.021). This effect was preserved after adjusting for AD biomarkers and their interactions with age in model 4B (*β*_synSAA×age_ = - 0.23 seconds per year, p=0.026; at age 60, 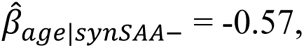 CI = -0.69 - -0.44; 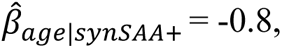 CI = -1.01 - -0.58). In addition, in Model 3B, faster age-related declines were seen in A+ vs A- (*β*_synSAA×age_ = -0.21 points per year, p=0.0039). Taken together, synSAA-/A- individuals showed the least decline per year on this test, while synSAA+/A+ individuals declined the fastest (at age 60, 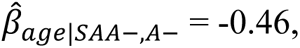 CI = -0.59 - -0.33; 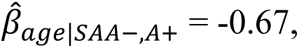 CI = -0.83 - -0.52; 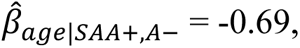 CI = -0.91 - -0.47; 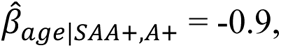 CI = -1.14 - -0.67).

#### PACC-3

No significant differences in age-related change are seen by synSAA status in either model 5A or 5B. In model 5B, faster declines are seen for 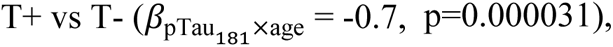 but not for A+ vs A-.

In an exploratory analysis, we reparametrized the above models to use binary pTau_181_/A*β*_42_ as a single indicator of AD biomarker positivity, and created a four-level variable representing combined status on this variable with synSAA status. As in previous models, we tested the main effects of this variable and its interactions with both age terms. The patterns were largely similar. For Trails B (Model 1C), the four-level interaction term with quadratic age was significant, so the full age structure was retained (4way × age^2^, p=0.017). Pairwise contrasts of the instantaneous age trends at selected ages indicated differences between the synSAA-/AD-group and the synSAA-/AD+ and synSAA+/AD+ groups, but only at older ages. Findings for the Trails difference score (Model 2C) were very similar. Digit Span Backward results indicated no significant interactions with age (Model 3C). For Digit Symbol, only the linear age interaction was significant (4way × age, p=0.0073), and pairwise trend contrasts once more indicated differences between the synSAA-/AD- group and the synSAA-/AD+ and synSAA+/AD+ groups (Model 4C), although after adjustment for multiplicity these did not reach statistical significance (.05 < p < .10). Finally, for PACC-3, again only the linear age interaction was significant (4way × age, p=0.0000050), and pairwise trend contrasts revealed differences between the synSAA-/AD+ group and the synSAA-/AD- and synSAA+/AD-groups (Model 5C). Full results of this analysis, including model summaries and simple slope contrasts, are shown in Supplementary Table 2 and Supplementary Figure 2.

## 4 Discussion

Results of the present study of a cohort of predominantly cognitively unimpaired, late-middle-aged participants showed syn, amyloid, and tau prevalances of 12%, 31%, and 21%, respectively. Observed co-occurrence of these markers suggests significant syn and AD copathology, specifically based on pTau_181_, but this relationship attenuates in magnitude and significance after adjusting for age. Several other cohorts have recently shown evidence of copathology, but the specifics have differed. In a cohort representing multiple etiologies, Bellomo and colleagues found increased occurrence of syn-seeds with increasing progression along the AD clinical continuum, even after adjusting for age^8^. Two other cohorts have established prevalent synSAA copathology with amyloid, but not tau. In BioFINDER, syn prevalence was higher among unimpaired, A+ individuals with and without adjusting for age^6^, but among impaired participants, no significant syn and AD copathology was found after adjusting for age, although a marginal link between syn and tau was observed^7^. However, the T marker used in BioFINDER was, variously, CSF pTau_217_ or tau PET, whereas the present analysis used only CSF pTau_181_. More recently, a study in ADNI relating synSAA to CSF A*β*_42_ and pTau suggested higher prevalence of syn among A+, but *lower* prevalence among T+^5^, in both impaired and unimpaired cohorts. This study also explored the frequency of AD neuropathological correlates in a subset that were positive for alpha synuclein at autopsy, but the relationship of Lewy bodies to A and T specifically was not reported. Taken together, emerging work suggests that syn copathology occurs as aged individuals progress along the AD clinical and biomarker spectrum. Further longitudinal follow-up will be needed to understand whether co-occurrence of AD and syn pathology reflects a mechanistic connection or is an epiphenomenon of age-related processes, and to explore the prognostic value of such copathology for development of Lewy body dementia.

In addition to synSAA and AD biomarkers, our deeply phenotyped cohort also had NTK results, allowing us to examine the effects of joint pathophysiology on these exploratory outcomes. Significant group differences were seen across markers, but the pattern of differences related most directly to T, with pairwise contrasts between groups that differed only on synSAA status generally proving nonsignificant. Across all NTK markers, mean levels were highest in the synSAA+/T+ group. To our knowledge this is the first study of NTK markers in a cohort characterized for syn, and future work will need to replicate this finding.

Relationships between the synSAA and cognitive status as adjudicated by clinical consensus suggest that the assay may be predictive of overall impairment in this population, which also agrees broadly with other recent work. However, we did not see strong relationships with specific neurological findings related to Lewy body disease. Findings from BioFINDER were somewhat similar to ours. In those without dementia, no relationship was observed between synSAA and motor symptoms^6^. Among memory clinic patients with MCI or dementia, a relationship between synSAA and motor symptoms was seen, but only among those who were synSAA+/AD-^7^. Similarly, in ADNI, no relationship was seen between synSAA and either sleep difficulties or hallucinations^5^. In our largely middle-aged sample that was targeted to be enriched for AD risk specifically, statistical power to detect co-ocurrence of syn-seeds with relatively rare LBD signs and symptoms was low. Additionally, clinical exams were performed by nurse practitioners rather than neurologists, and did not include full assessments for Parkinson’s disease and related disorders, and this imprecision may have reduced our assessment accuracy and statistical power further.

We observed steeper age-related decreases in Digit Symbol performance among synSAA+ individuals. Additionally, trajectories on both Trails B and the Trails Difference score worsened slightly faster among synSAA+ individuals, although this difference was marginal and attenuated to non-significant once AD biomarkers were added to the model. These findings were not corrected for multiple comparisons across outcomes, and should be interpreted with caution. However, if replicated, it would suggest that preclinical cognitive change, analogous to Stage 2 decline in the A/T/N framework for Alzheimer’s disease, can be observed in the Lewy body disease spectrum as well, on executive function tasks expected to show early change. Palmqvist and colleagues likewise found evidence for preclinical cognitive change associated with syn-seeds, but in contrast to the present work, this difference was observed across all cognitive outcomes^6^. Tosun and colleagues found evidence of steeper decline among AD+/SAA+ only on global assessments including ADAS-Cog and PACC, but not on executive function^5^. Differences between these studies may be due to the fact that both BioFINDER and ADNI have larger cohorts, or to differences in assessments; in particular, BioFINDER’s assessment of executive function made use of a test similar to our Digit Symbol task, but other assessments were different in substantive ways.

The study has some limitations that should be noted. First, our sample is a largely White convenience sample that has been upweighted for AD risk. Additional studies are needed in population-based samples. Second, because LPs were not generally available at the same time as the cognitive baseline, we have relatively few years of cognitive follow-up after the biomarker measurement, making a prospective design impractical. The design we chose instead was to use status at the most recent available LP to predict the full longitudinal trajectory, a largely retrospective design, which has inferential drawbacks. Third, because the original focus of the cohort studies was AD, our clinical assessments were not designed with Lewy body disorders in mind. Both cohorts are now adding the Uniform Parkinson’s Disease Rating Scale and the University of Pennsylvania Smell Identification Test to the portfolio of assessments in the hopes of getting a clearer clinical picture of this dementia etiology in our samples.

In summary, this study provides additional insight into the co-occurrence of syn and AD pathology. The utility of a qualitative seed amplification assay for syn pathology is apparent and expected to significantly enhance the ability to understand the contribution of multiple proteinopathies to cognitive decline and mixed dementia. Development of methods to measure proteinopathies in vivo (and early) will provide greater precision for preventing and treating dementia.

## Data Availability

All data used in the present study are available upon reasonable request. Interested parties should submit a resource request at https://wrap.wisc.edu/data-requests-2/.

## 5 Acknowledgements

The NeuroToolKit is a panel of exploratory prototype assays designed to robustly evaluate biomarkers associated with key pathologic events characteristic of AD and other neurological disorders, used for research purposes only and not approved for clinical use (Roche Diagnostics International Ltd, Rotkreuz, Switzerland). Elecsys β-amyloid(1–42) CSF and Elecsys Phospho-Tau (181P) CSF assays are approved for clinical use. COBAS and ELECSYS are trademarks of Roche. All other product names and trademarks are the property of their respective owners.

We are grateful to Rachel Weinberg and Amy Hawley, who each helped tremendously in facilitating this collaboration. We extend our deepest thanks to the WRAP and WADRC participants and staff for their invaluable contributions to the study.

## 5.1 Consent statement

All participants provided informed consent.

