## Supplement for "Misfolded alpha synuclein co-occurrence with Alzheimer’s disease proteinopathy"

### 1 Supplementary Table 1

Association between synSAA and variables of interest overall; stratified by age; and covarying age (Age-adjusted). For all analyses, assay results of "Detected-1" were coded as synSAA+, "Not Detected" as synSAA-, and all others as missing values. Primary analysis: no age adjustment computed; Sensitivity 1: Stratified analysis within younger participants (< 65) and older participants (≥ 65); Sensitivity 2: Age-adjusted general linear model (Age-adjusted) predicting synSAA positivity as a function of variables of interest after covarying age. For Primary and Sensitivity 1, p-values reflect chi-square tests of association; for Sensitivity 2, p-values reflect likelihood ratio tests comparing the full model to one containing only an age term.

| Variable | Age group | Risk group | Proportion synSAA+ | p |
|---|---|---|---|---|
| $A\beta_{42/40}$ positivity | Overall | A- | 29/285 (10%; 7%-14%) | 0.17 |
| | | A+ | 19/127 (15%; 10%-22%) | |
| | < 65 | A- | 13/164 (8%; 5%-13%) | 0.49 |
| | | A+ | 1/29 (3%; -1%-19%) | |
| | ≥ 65 | A- | 16/121 (13%; 8%-21%) | 0.36 |
| | | A+ | 18/98 (18%; 12%-27%) | |
| | Age-adjusted | A- | – (10%; 7%-15%) | 0.90 |
| | | A+ | – (11%; 6%-18%) | |
| $pTau_{181}/A\beta_{42}$ positivity | Overall | AD- | 27/287 (9%; 7%-13%) | 0.067 |
| | | AD+ | 19/118 (16%; 10%-24%) | |
| | < 65 | AD- | 13/169 (8%; 4%-13%) | 0.71 |
| | | AD+ | 1/22 (5%; -1%-24%) | |
| | ≥ 65 | AD- | 14/118 (12%; 7%-19%) | 0.18 |
| | | AD+ | 18/96 (19%; 12%-28%) | |
| | Age-adjusted | AD- | – (10%; 7%-14%) | 0.56 |
| | | AD+ | – (12%; 7%-19%) | |

| Variable | Age group | Risk group | Proportion synSAA+ | p |
|---|---|---|---|---|
| $pTau_{181}$ positivity | Overall | T- | 30/322 (9%; 7%-13%) | 0.011 |
| | | T+ | 17/88 (19%; 12%-29%) | |
| | < 65 | T- | 12/176 (7%; 4%-12%) | 0.17 |
| | | T+ | 3/19 (16%; 5%-38%) | |
| | ≥ 65 | T- | 18/146 (12%; 8%-19%) | 0.15 |
| | | T+ | 14/69 (20%; 12%-31%) | |
| | Age-adjusted | T- | – (9%; 7%-13%) | 0.14 |
| | | T+ | – (15%; 9%-24%) | |
| Cognitive status | Overall | CU | 39/375 (10%; 8%-14%) | 0.00050 |
| | | MCI | 5/30 (17%; 7%-34%) | |
| | | Dementia | 5/6 (83%; 42%-99%) | |
| | < 65 | CU | 14/192 (7%; 4%-12%) | 0.14 |
| | | MCI | 1/2 (50%; 9%-91%) | |
| | | Dementia | 0/0 (NaN%; 0%-100%) | |
| | ≥ 65 | CU | 25/183 (14%; 9%-19%) | 0.00100 |
| | | MCI | 4/28 (14%; 5%-32%) | |
| | | Dementia | 5/6 (83%; 42%-99%) | |
| | Age-adjusted | CU | – (10%; 7%-14%) | 0.0024 |
| | | MCI | – (10%; 4%-25%) | |
| | | Dementia | – (75%; 25%-96%) | |
| Neurological findings | Overall | Normal | 31/311 (10%; 7%-14%) | 0.018 |
| | | Age-related | 8/72 (11%; 5%-21%) | |
| | | Dementing | 8/29 (28%; 15%-46%) | |
| | < 65 | Normal | 8/161 (5%; 2%-10%) | 0.035 |
| | | Age-related | 3/25 (12%; 3%-31%) | |
| | | Dementing | 2/8 (25%; 6%-60%) | |
| | ≥ 65 | Normal | 23/150 (15%; 10%-22%) | 0.16 |
| | | Age-related | 5/47 (11%; 4%-23%) | |
| | | Dementing | 6/21 (29%; 14%-50%) | |
| | Age-adjusted | Normal | – (10%; 7%-14%) | 0.27 |
| | | Age-related | – (9%; 4%-17%) | |

| Variable | Age group | Risk group | Proportion synSAA+ | p |
|---|---|---|---|---|
| | | Dementing | – (19%; 9%-37%) | |
| Parkinsonian signs | Overall | Absent | 44/395 (11%; 8%-15%) | 0.16 |
| | | Present | 5/22 (23%; 10%-44%) | |
| | < 65 | Absent | 15/192 (8%; 5%-13%) | 1.00 |
| | | Present | 0/5 (0%; -5%-49%) | |
| | ≥ 65 | Absent | 29/203 (14%; 10%-20%) | 0.15 |
| | | Present | 5/17 (29%; 13%-53%) | |
| | Age-adjusted | Absent | – (10%; 8%-14%) | 0.54 |
| | | Present | – (14%; 5%-33%) | |
| REM behavior disorder | Overall | Absent | 47/411 (11%; 9%-15%) | 0.15 |
| | | Present | 2/6 (33%; 9%-70%) | |
| | < 65 | Absent | 14/194 (7%; 4%-12%) | 0.22 |
| | | Present | 1/3 (33%; 6%-80%) | |
| | ≥ 65 | Absent | 33/217 (15%; 11%-21%) | 0.39 |
| | | Present | 1/3 (33%; 6%-80%) | |
| | Age-adjusted | Absent | – (10%; 8%-14%) | 0.20 |
| | | Present | – (29%; 7%-69%) | |
| New-onset depression | Overall | Absent | 46/404 (11%; 9%-15%) | 0.40 |
| | | Present | 3/13 (23%; 7%-51%) | |
| | < 65 | Absent | 13/192 (7%; 4%-11%) | 0.052 |
| | | Present | 2/5 (40%; 12%-77%) | |
| | ≥ 65 | Absent | 33/212 (16%; 11%-21%) | 1.00 |
| | | Present | 1/8 (12%; 0%-49%) | |
| | Age-adjusted | Absent | – (10%; 8%-14%) | 0.22 |
| | | Present | – (22%; 7%-51%) | |

### 2 Supplementary Table 2

Results of exploratory nested linear mixed effects models of cognitive tests associated with executive function (Trail-Making Test B; Trail-Making Test B-A Difference Score; Digit Span Backward; Digit Symbol Substitution Test) and a global preclinical Alzheimer's cognitive composite (PACC-3). Model set C included a single-dimensional measure of Alzheimer's pathology, $pTau_{181}/A\beta_{42}$, and its interactions with age and age$^2$. Nonsignificant interaction terms (p>.1) were removed.

#### *2.1 Post-hoc tests*

To explore significant interactions between age and biomarker group (synSAA-/AD-, synSAA-/AD+, synSAA+/AD-, synSAA+/AD+) suggestive of group differences in cognitive change, we conducted pairwise comparisons between simple age-related slopes. Holm-Bonferroni corrections were applied to control familywise type I error.

Pairwise contrasts:

##### *2.1.1 Trail-Making Test B*

| contrast | age_c60 | estimate | SE | df | t.ratio | p.value |
|---|---|---|---|---|---|---|
| (synSAA-/AD-) - (synSAA-/AD+) | -10 | 0.653 | 0.551 | 562 | 1.185 | 0.63659 |
| (synSAA-/AD-) - (synSAA+/AD-) | -10 | -0.226 | 0.915 | 430 | -0.2471 | 0.99469 |
| (synSAA-/AD-) - (synSAA+/AD+) | -10 | 0.8092 | 1.082 | 595 | 0.7479 | 0.87754 |
| (synSAA-/AD+) - (synSAA+/AD-) | -10 | -0.879 | 1.004 | 455 | -0.8756 | 0.8175 |
| (synSAA-/AD+) - (synSAA+/AD+) | -10 | 0.1563 | 1.155 | 604 | 0.1353 | 0.99911 |
| (synSAA+/AD-) - (synSAA+/AD+) | -10 | 1.0353 | 1.369 | 519 | 0.7561 | 0.87406 |
| (synSAA-/AD-) - (synSAA-/AD+) | 0 | -0.2641 | 0.302 | 334 | -0.8735 | 0.81856 |
| (synSAA-/AD-) - (synSAA+/AD-) | 0 | -0.3105 | 0.498 | 323 | -0.6235 | 0.92449 |
| (synSAA-/AD-) - (synSAA+/AD+) | 0 | -0.6518 | 0.637 | 355 | -1.0226 | 0.73633 |
| (synSAA-/AD+) - (synSAA+/AD-) | 0 | -0.0464 | 0.54 | 325 | -0.0859 | 0.99977 |

| contrast | age_c60 | estimate | SE | df | t.ratio | p.value |
|---|---|---|---|---|---|---|
| (synSAA-/AD+) - (synSAA+/AD+) | 0 | -0.3877 | 0.669 | 353 | -0.5799 | 0.93808 |
| (synSAA+/AD-) - (synSAA+/AD+) | 0 | -0.3413 | 0.779 | 345 | -0.438 | 0.97186 |
| (synSAA-/AD-) - (synSAA-/AD+) | 10 | -1.1812 | 0.396 | 472 | -2.9847 | 0.01575 |
| (synSAA-/AD-) - (synSAA+/AD-) | 10 | -0.3949 | 0.757 | 488 | -0.5216 | 0.95389 |
| (synSAA-/AD-) - (synSAA+/AD+) | 10 | -2.1128 | 0.615 | 362 | -3.4361 | 0.00367 |
| (synSAA-/AD+) - (synSAA+/AD-) | 10 | 0.7863 | 0.769 | 478 | 1.0219 | 0.7367 |
| (synSAA-/AD+) - (synSAA+/AD+) | 10 | -0.9316 | 0.63 | 356 | -1.4777 | 0.4521 |
| (synSAA+/AD-) - (synSAA+/AD+) | 10 | -1.7179 | 0.903 | 421 | -1.9029 | 0.22836 |

#### *2.1.2 Trail-Making Test B-A Difference Score*

| contrast | age_c60 | estimate | SE | df | t.ratio | p.value |
|---|---|---|---|---|---|---|
| (synSAA-/AD-) - (synSAA-/AD+) | -10 | 0.4512 | 0.504 | 530 | 0.8955 | 0.80721 |
| (synSAA-/AD-) - (synSAA+/AD-) | -10 | -0.2975 | 0.829 | 368 | -0.359 | 0.98414 |
| (synSAA-/AD-) - (synSAA+/AD+) | -10 | 0.4764 | 0.987 | 681 | 0.4825 | 0.96296 |
| (synSAA-/AD+) - (synSAA+/AD-) | -10 | -0.7488 | 0.91 | 396 | -0.8224 | 0.84385 |
| (synSAA-/AD+) - (synSAA+/AD+) | -10 | 0.0252 | 1.054 | 677 | 0.0239 | 1 |
| (synSAA+/AD-) - (synSAA+/AD+) | -10 | 0.774 | 1.245 | 523 | 0.6218 | 0.92509 |
| (synSAA-/AD-) - (synSAA-/AD+) | 0 | -0.2066 | 0.278 | 333 | -0.7444 | 0.87903 |
| (synSAA-/AD-) - (synSAA+/AD-) | 0 | -0.0735 | 0.458 | 318 | -0.1605 | 0.99853 |
| (synSAA-/AD-) - (synSAA+/AD+) | 0 | -0.8183 | 0.579 | 384 | -1.4135 | 0.49165 |
| (synSAA-/AD+) - (synSAA+/AD-) | 0 | 0.1331 | 0.495 | 319 | 0.2687 | 0.9932 |
| (synSAA-/AD+) - (synSAA+/AD+) | 0 | -0.6117 | 0.607 | 377 | -1.0084 | 0.74463 |
| (synSAA+/AD-) - (synSAA+/AD+) | 0 | -0.7447 | 0.71 | 358 | -1.0489 | 0.72073 |
| (synSAA-/AD-) - (synSAA-/AD+) | 10 | -0.8645 | 0.376 | 499 | -2.2972 | 0.10004 |
| (synSAA-/AD-) - (synSAA+/AD-) | 10 | 0.1505 | 0.721 | 505 | 0.2087 | 0.99678 |
| (synSAA-/AD-) - (synSAA+/AD+) | 10 | -2.113 | 0.575 | 380 | -3.6749 | 0.00155 |
| (synSAA-/AD+) - (synSAA+/AD-) | 10 | 1.0149 | 0.732 | 496 | 1.3862 | 0.50858 |
| (synSAA-/AD+) - (synSAA+/AD+) | 10 | -1.2485 | 0.589 | 374 | -2.1188 | 0.14898 |
| (synSAA+/AD-) - (synSAA+/AD+) | 10 | -2.2634 | 0.852 | 438 | -2.6558 | 0.04074 |

#### *2.1.3 Digit Symbol*

| contrast | estimate | SE | df | t.ratio | p.value |
|---|---|---|---|---|---|
| (synSAA-/AD-) - (synSAA-/AD+) | 0.2072 | 0.0808 | 282 | 2.566 | 0.0525 |
| (synSAA-/AD-) - (synSAA+/AD-) | 0.2634 | 0.1364 | 310 | 1.932 | 0.217 |
| (synSAA-/AD-) - (synSAA+/AD+) | 0.3786 | 0.1572 | 258 | 2.408 | 0.0781 |
| (synSAA-/AD+) - (synSAA+/AD-) | 0.0562 | 0.1445 | 302 | 0.389 | 0.98 |
| (synSAA-/AD+) - (synSAA+/AD+) | 0.1714 | 0.1606 | 253 | 1.068 | 0.7094 |
| (synSAA+/AD-) - (synSAA+/AD+) | 0.1152 | 0.1974 | 277 | 0.584 | 0.9369 |

#### *2.1.4 PACC-3*

| contrast | estimate | SE | df | t.ratio | p.value |
|---|---|---|---|---|---|
| (synSAA-/AD-) - (synSAA-/AD+) | 0.4644 | 0.102 | 2374 | 4.552 | 0.000033 |
| (synSAA-/AD-) - (synSAA+/AD-) | -0.0715 | 0.186 | 2343 | -0.384 | 0.980725 |
| (synSAA-/AD-) - (synSAA+/AD+) | 0.6019 | 0.191 | 2362 | 3.159 | 0.008697 |
| (synSAA-/AD+) - (synSAA+/AD-) | -0.5359 | 0.196 | 2359 | -2.731 | 0.03228 |
| (synSAA-/AD+) - (synSAA+/AD+) | 0.1375 | 0.196 | 2344 | 0.702 | 0.89614 |
| (synSAA+/AD-) - (synSAA+/AD+) | 0.6734 | 0.253 | 2346 | 2.659 | 0.039413 |

### 4 Supplementary Figure 1

Forest plots showing proportions of people in groups of interest with positive synSAA status, overall and stratified by age (<65 vs >=65). Left: Biomarker groupings (A=A$\beta_{42}$; AD=pTau$_{181}$/A$\beta_{42}$; T=pTau$_{181}$); Right: Cognitive signs and symptoms.

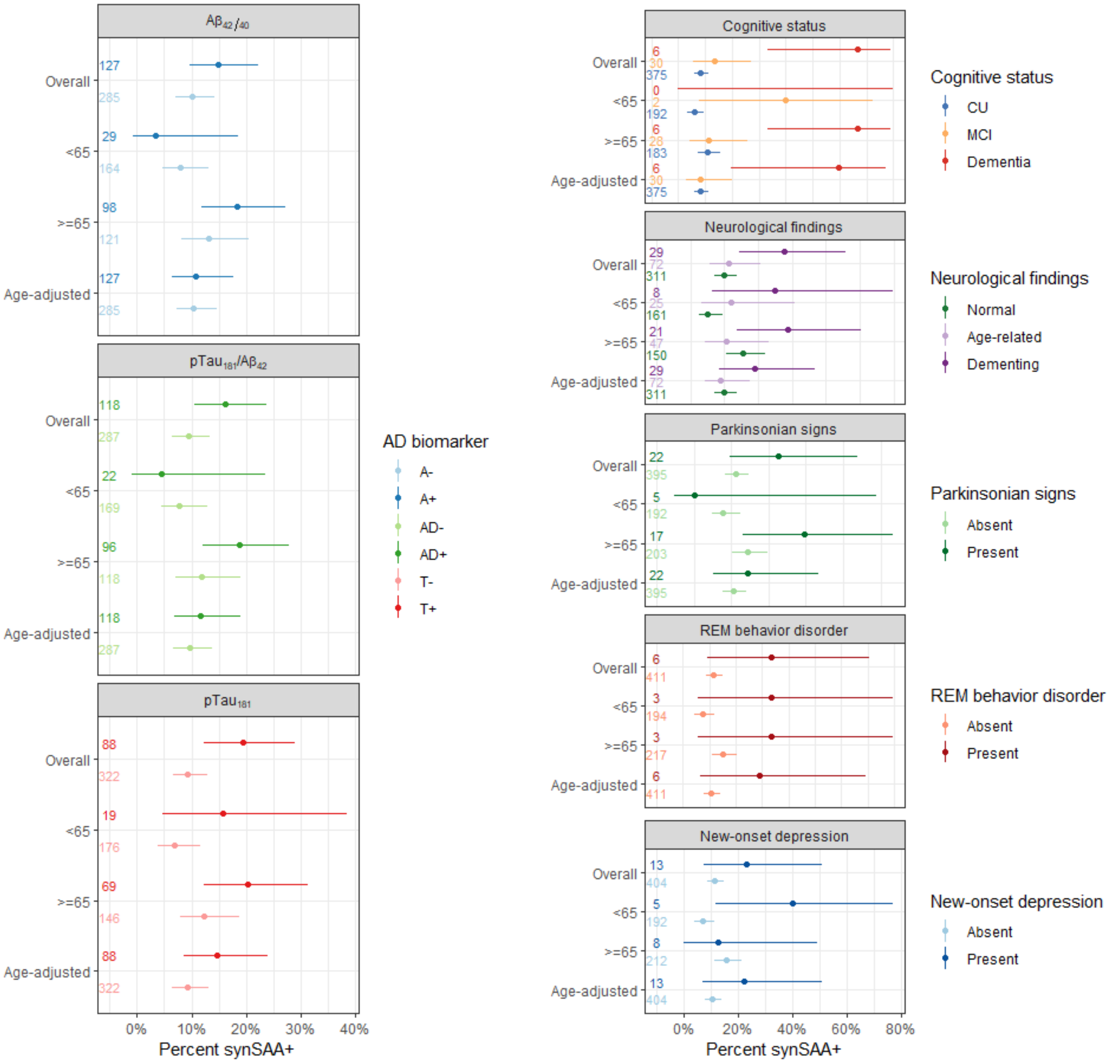

Aβ42/40
pTau181/Aβ42
pTau181
Overall
<65
>=65
Age-adjusted
Percent synSAA+
AD biomarker
A-
A+
AD-
AD+
T-
T+
Cognitive status
CU
MCI
Dementia
Neurological findings
Normal
Age-related
Dementing
Parkinsonian signs
Absent
Present
REM behavior disorder
Absent
Present
New-onset depression
Absent
Present

### 5 Supplementary Figure 2

Results of exploratory nested linear mixed effects models of cognitive tests associated with executive function (Trail-Making Test B; Digit Span Backward; Digit Symbol Substitution Test) and a global preclinical Alzheimer's cognitive composite (PACC-3). Model-predicted values and confidence bands derived from final models C represented in Supplementary Table 2. Predictors not shown directly in the graph have been set to their average value. The largest model examined the effect of binary synSAA, synSAA $\times$ age, and synSAA $\times$ age$^2$, controlling for sex, education, and prior exposure to the battery. Unlike the models illustrated in Figure 1, these models include $\text{pTau}_{181}/\text{A}\beta_{42}$ as a unidimensional measure of AD pathophysiology, along with its interactions with age, age$^2$, and synSAA. From this largest model, nonsignificant interaction terms ($p>.1$) were removed. The spaghetti plot layer beneath represents individual participants' measurements over time.

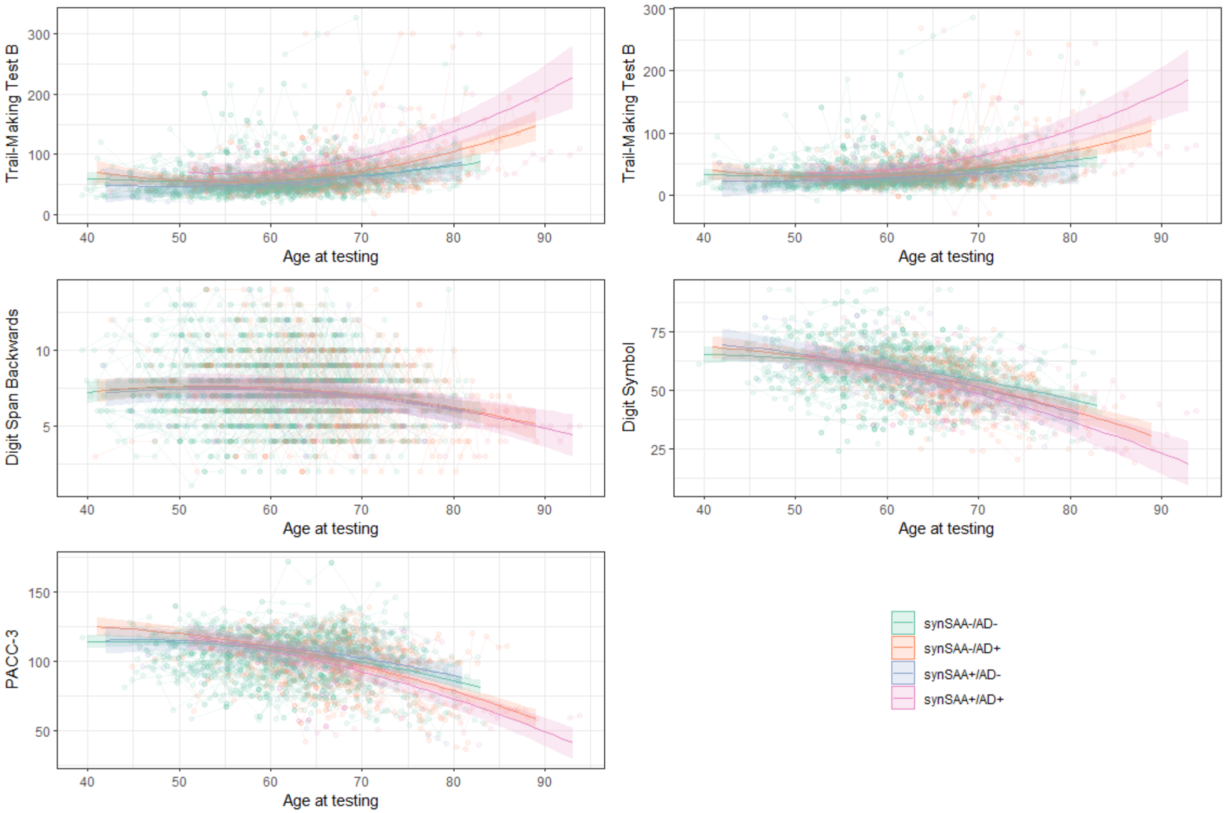

Trail-Making Test B
Trail-Making Test B
Digit Span Backwards
Digit Symbol
PACC-3
Age at testing
synSAA-/AD-
synSAA-/AD+
synSAA+/AD-
synSAA+/AD+
